# Understanding the genetic epidemiology of hereditary breast cancer in India using whole genome data from 1029 healthy individuals

**DOI:** 10.1101/2023.10.20.23297296

**Authors:** Aastha Vatsyayan, Prerika Mathur, Rahul C Bhoyar, Mohamed Imran, Vigneshwar Senthivel, Mohit Kumar Divakar, Anushree Mishra, Bani Jolly, Sridhar Sivasubbu, Vinod Scaria

**Affiliations:** CSIR Institute of Genomics and Integrative Biology (CSIR-IGIB) Mathura Road, Delhi 110025, India; Academy of Scientific and Innovative Research (AcSIR), Ghaziabad 201002, India; Department of Biological Sciences, Birla Institute of Technology and Science, Pilani, Pilani Campus, Rajasthan, India; Vishwanath Cancer Care Foundation, B 702, Neelkanth Business Park Kirol Village, Mumbai, 400 086, India

## Abstract

**Aim:** Breast cancer is the most highly reported cancer in India as well as globally (Globocan 2020). Genetic testing could help tackle the increasing cancer burden by enabling carriers of pathogenic variants obtain an early diagnosis through increased surveillance, and help guide treatment, and in some cases enable prevention. However, accurate interpretation of variant pathogenicity must be established in a population-specific manner to ensure effective use of genetic testing. Further, since *BRCA1* and *BRCA2* are importance breast cancer genes, even rare variants must be studied for their potential effect on the disease.

**Materials and Methods:** We query the IndiGen data obtained from whole genome sequencing of 1029 Indian individuals, and perform variant classification of all reported BRCA variants according to the gold-standard ACMG/AMP guidelines to establish disease epidemiology. We further implement machine learning approaches to classify all reported non-benign variants, and create a ready-reference of variants classified in a manner close to ACMG guidelines at scale.

**Results:** We determined the genetic prevalence to be the following: 1 in nearly 341 individuals for *BRCA1*, and 1 in nearly 170 individuals for *BRCA2* are likely to be carriers of pathogenic mutations. Overall, 1 in nearly 114 individuals are likely to be carriers of pathogenic BRCA mutations. Further, using the brca-NOVUS tool, we classified 1,54,045 genetic variants across 18 population sets and 4 large variant repositories as either pathogenic or benign.

**Conclusion:** The high population prevalence indicates a greater need of studying genetic variants linked with breast cancer in an Indian population specific manner. To the best of our knowledge, this is the first and most comprehensive population-scale genetic epidemiological study of BRCA-linked breast cancer variants reported from India.

## Background

Cancer is a complex genetic disorder encompassing more than 277 types of disease^1^. It is a leading cause of death globally, with Asia leading in both incidence and mortality numbers^2^. India bears a rapidly growing cancer burden as well: from an estimated 1.46 million cases in 2022, cancer incidence is expected to have reached 1.57 million cases in 2025^3^. Breast cancer recorded the highest number of new cases in 2020 both globally (11.7%) as well as in India (13.5%)^2^, making it a major cause for concern. In India, 1 in 29 women are likely to develop breast cancer^4^. Further, 1 in 22 women in urban and 1 in 60 women in rural areas are likely to develop breast cancer during their lifetime^5^.

About 5 - 10% of all cancers are hereditary in nature and form a part of familial or hereditary cancer syndromes. Each of these syndromes confer an increased lifetime risk to a certain group of cancers on the bearer of related pathogenic germline mutations. Although the number seems small, studying pathogenic variants associated with familial cancers could significantly benefit patients with early diagnosis through increased surveillance, guiding treatment, and in some cases prevention. Since these cancers are caused due to heritable genetic variants, a number of individuals in the family and community (such as the Ashkenazi Jewish community) could extensively benefit from genetic screening.

Understanding the importance of genetic screening, the National Comprehensive Cancer Network (NCCN)^6^ guidelines have incorporated it as part of their guidelines for detection, prevention and risk reduction in breast cancer. A person with a known personal or family history of any of the cancers linked with Hereditary Breast and Ovarian Cancer (HBOC) is eligible to undergo genetic testing. However, a bottleneck still remains in form of accurate interpretation of the testing results: a large number of variants remain classified as variants of uncertain significance (VUS), while some others may be misclassified due to lack of implementation of uniform interpretation guidelines. To address this lacuna, the American College of Medical Genetics and Genomics (ACMG) along with the Association for Molecular Pathology (AMP) have issued a set of gold-standard guidelines^7^ for the accurate classification of sequence variants into five categories: Pathogenic, Likely Pathogenic, Benign, Likely Benign and Variant of Uncertain Significance (VUS). The guidelines take into account 28 attributes that consider evidence spanning across population allele frequencies, pathogenicity prediction scores, annotations from benchmark databases, as well as exhaustive literature review to offer an accurate classification based on all current information available.

Another largely unaddressed obstacle preventing effective genetic testing is the lack of variant studies in a population-specific manner. As a consequence of factors such as migration, selection and genetic drift^8^, different populations bear a different prevalence of causative variants, and in some cases variants unique to a given population. This necessitates the establishment of the variant pathogenicity landscape of a disease-linked gene in a population-specific manner. However, large-scale population datasets such as gnomAD^9^ and 1000 Genomes^10^ (1KG) do not represent Indian populations sufficiently. For example in the gnomAD database, South Asian populations form only about 3.2% of their total data. The creation of the IndiGen^11^ dataset, a whole genome sequencing project from 1029 self-declared healthy Indian individuals from 27 states, enables us to study HBOC variants in the context of Indian populations and subpopulations.

In this study, we use the IndiGen data to establish the variant landscape using ACMG/AMP guidelines to classify all variants reported in the *BRCA1* and *BRCA2* genes linked with HBOC. We have analyzed the pathogenic variants thus obtained to establish the genetic epidemiology of BRCA-linked HBOC in India. Finally, given the very large number of variants of unknown or uncertain significance (VUS) across the BRCA genes, we have employed machine learning approaches to classify variants based on the ACMG guidelines. Through the brca-NOVUS tool, which is trained on a ACMG/AMP classified variant dataset, we have classified all VUS reported in IndiGen, as well as 18 other population-scale datasets. We have also classified all non-benign variants obtained from four large scale BRCA variant repositories, thus effectively establishing a repository that can act as a ready-reference for dealing with VUS in clinical and research settings.

To the best of our knowledge, this is the first and most comprehensive population-scale genetic epidemiological study of BRCA-linked breast cancer variants reported from India.

## Materials and Methods

### IndiGen dataset and variant annotation

The IndiGen dataset was derived from the whole genome sequencing of 1029 cosmopolitan self-declared healthy Indians across 27 states; well-written informed consent was obtained. We queried 59,646,267 genetic variants including single nucleotide variants and Indels across the dataset for 2 high risk breast cancer genes associated with breast cancer^12^ *-BRCA1* and *BRCA2*. The variants were annotated using ANNOVAR^13^ (ver 2018-04-06), which annotated the variants with data from RefGene^14^, and dbSNP^15^ that describe a variant, dbNSFP35a^16^ that offers various pathogenicity prediction scores, as well as allele frequencies from global population datasets including gnomAD v3, 1KG, Esp6500^17^ and Greater Middle East (GME)^18^. The clinical significance of each variant was obtained by programmatically querying the ClinVar^19^ (ver 2020-01-13) database using custom scripts.

### ACMG/AMP classification and establishment of genetic epidemiology

The BRCA variants obtained from IndiGen were classified using the ACMG/AMP guidelines. Briefly, each variant was assessed for 28 attributes that spanned population allele frequencies, computational prediction, annotations from benchmarked databases, as well as exhaustive literature survey. The final classification was obtained based on the attributes marked using the Genetic Variant Interpretation Tool^20^. Details of the guidelines used for annotation of attributes are provided in Supplementary Data 1. Based on the pathogenic variants obtained, the genetic epidemiology was calculated for each gene.

### Comparison with other populations

#### Pathogenic IndiGen variants in global populations

The pathogenic variants so obtained were searched across 19 global population datasets for allele frequencies including GnomAD populations and subpopulations, the China Metabolic Analytics Project (ChinaMAP)^21^, the Hong Kong Cantonese population (HKG)^22^, TogoVar (Japanese population)^23^, Korea1K Variome (KGP)^24^, Korean Variant Archive 2 (KOVA 2)^25^, Qatar^26^, Taiwan Biobank^27^, GME populations and subpopulations, 1KG, the Gambian dataset^28^ (Gambian), the GenomeAsia100K Project^29^ (GenomeAsia), Human Genome Diversity Project (HGDP)^30^, the Andamanese population^31^(Andamanese), the Simons Genome Diversity Project^32^(Simons), the Singapore Sequencing Indian Project (SSIP)^33^, and the Singapore Sequencing Malay Project (SSMP)^34^. Additionally, the Vietnamese Genetic Variation Database^35^, and the Iranome^36^ were also queried.

### Population specific ACMG-AMP reclassification and comparison

In order to facilitate a better understanding of the prevalence of pathogenic BRCA variants, we performed the ACMG-AMP classification of all exonic *BRCA1* and *BRCA2* variants in the Qatari population^26^ that had been previously sequenced by Fakhro et. al. Whole genome and exome sequencing was performed for 1,376 individuals, and 20.9 million SNPs and 3.1 million indels were obtained.

### ML Classification of variants

Since a majority of IndiGen variants (80.2%) across both genes were of either unknown or uncertain significance (VUS) due to a lack of functional evidence, we decided to classify these variants using the brca-NOVUS tool, a previously standardized machine learning model developed for classification of exonic BRCA variants by our lab. The model has been trained on an ACMG-classified dataset of BRCA variants, and thus offers predictions close to ACMG-classification at scale.

We collected all variants reported across the following: (a) 18 population-scale datasets including IndiGen, gnomAD, ChinaMAP, HKG, TogoVar, KGP, KOVA 2, Qatar, Taiwan Biobank, GME populations and subpopulations, 1KG, Gambian, GenomeAsia, HGDP, Andamanese, Simons, SSIP, and SSMP.

(b) 4 large scale BRCA variant repositories including BRCAExchange^37^, Mastermind^38^, ARUP, as well as all BRCA ClinVar VUS variants.

Next, we removed all variants annotated by ClinVar as benign on likely benign. We processed the remaining variants from across all populations and each dataset, and ran our model on the resulting file.

## Results

### ACMG/AMP classification and establishment of genetic epidemiology

Upon querying the IndiGen data, we obtained a total of 3408 BRCA variants: 1906 of which belonged to *BRCA1* and 1502 to *BRCA2*. The annotated data was the carefully classified as per the guidelines detailed by ACMG/AMP. Finally, stringent quality control was applied where incorrectly called variants, or those with low quality (Read Depth<10 and Genotype Quality<20) were removed. Upon final tally, 3 pathogenic/likely pathogenic *BRCA1* and 5 *BRCA2* variants were obtained. Details of the variants and their ACMG attributes are given in Table 1 and summarized in Figure 1. A complete list of variants and their attributes is included in Supplementary Table 1.

**Table 1:** Table depicting the Pathogenic variants obtained after application of quality cut-offs and classification through ACMG guidelines.

| ID | Variant | Gene | Attributes | ACMG Classification |
| --- | --- | --- | --- | --- |
| 17:43071239:C:G | NM_007294:exon15:c.4676- | BRCA1 | PP5,PM2,PVS1 | Pathogenic (Ic) |
|  | 1G>C |  |  |  |
| 17:43091924:G:A | NM_007294:exon10:c.3607C>T:p.R1203X | *BRCA1* | PP5,PM2,PS3,PVS1 | Pathogenic (Ia) |
| 17:43093856:CTT:C | NM_007294:exon10:c.1673_1674del:p.K558Rfs*2 | *BRCA1* | PP5,PM2,PVS1 | Pathogenic (Ic) |
| 13:32336777:G:T | NM_000059:exon11:c.2422G>T:p.E808X | *BRCA2* | PVS1,PM2,PP3 | Pathogenic 1C |
| 13:32337156:GT:G | NM_000059:exon11:c.2802delT:p.D935Ifs*25 | *BRCA2* | PVS1,PM2 | Likely pathogenic (I) |
| 13:32338167:C:G | NM_000059:exon11:c.3812C>G:p.S1271X | *BRCA2* | PVS1, PP5, PP3 | Pathogenic (Id) |
| 13:32339502:AT:A | NM_000059:exon11:c.5148delT:p.Y1716* | *BRCA2* | PVS1,PM2 | Likely pathogenic (I) |
| 13:32370992:C:T | NM_000059:exon20:c.8524C>T:p.R2842C | *BRCA2* | PM1,PM2,PP3,PS3 | Likely pathogenic (II) |

**Figure 1:**
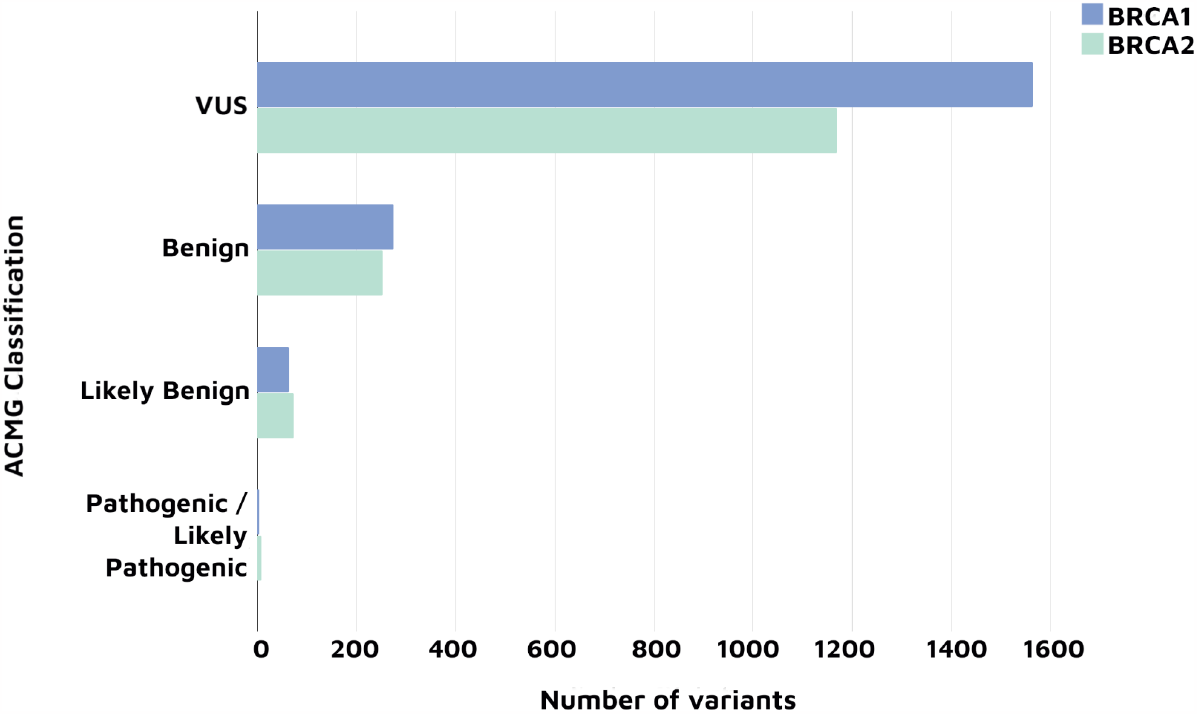
Figure summarizing the breakdown of the ACMG classification of *BRCA1* and *BRCA2* variants of the IndiGen data

The genetic prevalence was then established for each gene using the total number of pathogenic variants obtained, using the following formula:

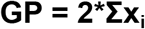

where, GP = Genetic Prevalence, x_i_ = allele frequency of pathogenic variants

We determined the genetic prevalence to be the following:

1 in nearly 341 individuals for *BRCA1*, and 1 in nearly 170 individuals for *BRCA2* are likely to be carriers of pathogenic mutations.

Overall, 1 in nearly 114 individuals are likely to be carriers of pathogenic BRCA mutations.

### Comparison with other populations

#### Pathogenic IndiGen variants in global populations

The 8 IndiGen pathogenic/likely pathogenic variants grepped across 19 population-scale datasets. However, only two variants found hits in the GnomAD database, indicating that these are rare variants largely specific to the Indian populations. The two variants include one *BRCA1* variant in the African/African American sub-population, and one *BRCA2* variant in the Latino/Admixed American. The variants are described in Table 2.

**Table 2:**
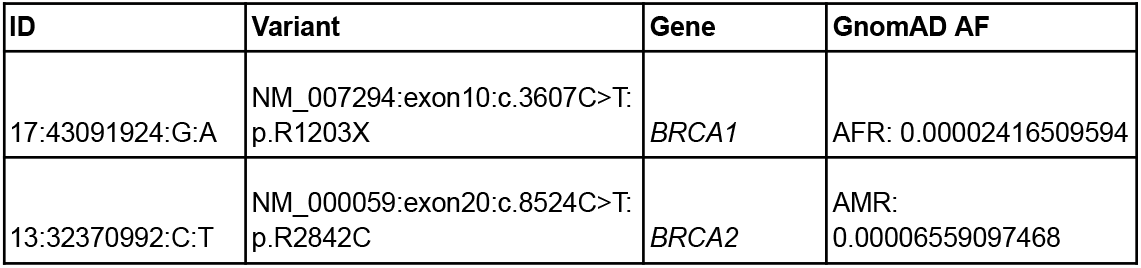
Table summarizing the IndiGen ACMG-classified pathogenic variants found in the GnomAD dataset.

| ID | Variant | Gene | GnomAD AF |
| --- | --- | --- | --- |
| 17:43091924:G:A | NM_007294:exon10:c.3607C>T:p.R1203X | *BRCA1* | AFR: 0.00002416509594 |
| 13:32370992:C:T | NM_000059:exon20:c.8524C>T:p.R2842C | *BRCA2* | AMR: 0.00006559097468 |

### Population specific ACMG-AMP reclassification and comparison

We ran the ANNOVAR tool on the Qatar data, and collected all recorded BRCA variants. We thus obtained a total of 918 BRCA variants, none of which were ClinVar pathogenic/likely pathogenic. We proceeded to reclassify all exonic variants that were not classified as benign/likely benign by ClinVar, and discovered 1 variant (rs786201716) to be Likely benign (I), while the others remained VUS due to lack of evidence.

### ML Classification of variants

Upon querying our 18 population-scale datasets, we obtained a total of 12,904 BRCA variants. After removing all variants reported by ClinVar as benign or likely benign, and running the model, we were left with 3,797 exonic variants for which we made population-wise predictions. Thus a total of 2908 unique exonic variants that were not benign were classified by our model across both genes. The cumulative allele frequencies (CAF) of pathogenic variants across each gene and each population are shown in Figure 2. The CAF are shown in Supplementary Table 2, while individual population-wise predictions are shown in Supplementary Table 3.

**Figure 2:**
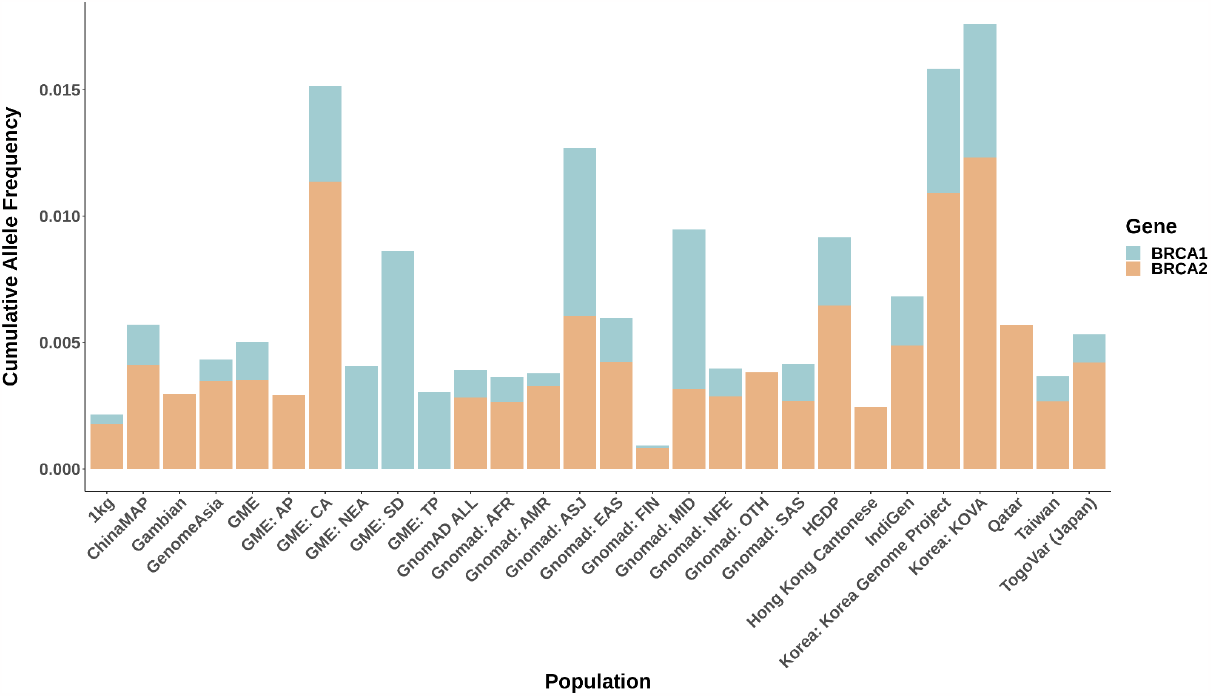
Plot depicting the cumulative allele frequencies of pathogenic variants across each population for both genes. **Abbreviations**: GnomAD - AFR: African/African American, AMI: Amish, AMR: Latino/Admixed American, ASJ: Ashkenazi Jewish, EAS: East Asian, FIN: Finnish, NFE: Non-Finnish European, MID: Middle Eastern, SAS: South Asian, OTH: Other (population not assigned) GME - NWA: Northwest Africa, NEA: Northeast Africa, AP: Arabian Peninsula, SD: Syrian Desert, TP: Turkish Peninsula, CA: Central Asia

We then collected a total of 2,29,814 variants from across 4 large variant repositories. After removing all ClinVar benign/likely benign variants, we were left with 1,93,336 variants across both genes. Upon running brca-NOVUS, we obtained a total of 1,50,248 unique exonic BRCA variants classified as either pathogenic or benign.

A complete list of all data repository variants classified along with their predictions is included in Supplementary Tables 4-7.

### Comparison of ML predictions with ACMG classification

#### IndiGen variants

A total of 99 exonic variants were classified by brca-NOVUS, of which 12 were predicted as pathogenic (3 *BRCA1* and 9 *BRCA2*). The tool was able to accurately identify all exonic variants classified as pathogenic through the application of ACMG guidelines and before removal from prevalence calculations due to quality control. It additionally classified one *BRCA2* variants as pathogenic: rs397507725 (p.I1516Dfs*13), which was classified as VUS due to lack of functional evidence through ACMG classification. The variant was assigned the PVS1-Very Strong attribute, since it is predicted to undergo Nonsense Mediated Decay (NMD) and is present in a biologically relevant transcript. It is also ClinVar Pathogenic and was thus assigned the PP5 attribute. It is a rare variant missing from all other populations. Additionally, it is also annotated as a high confidence loss-of-function variant by the Loftee tool. If more functional evidence becomes available, it is likely that the ACMG classification of the variant would be changed to pathogenic.

Thus, the brca-NOVUS predictions match closely with the ACMG classifications, with one variant identified as a target for further study.

#### Qatar variants

We compared the predictions generated for the Qatar data with the ACMG classifications performed, and discovered that the model predicted 21 exonic variants as benign, and 1 *BRCA2* variant as pathogenic - rs80358908 (G2281E) is an ACMG classified VUS variant largely missing from other populations (PM2), and is predicted to be pathogenic by multiple computational tools (PP3).

21 of the 22 variants were ACMG classified as VUS due to a lack of functional evidence. 1 variant was marked as benign by both ACMG and the model. Thus, through brca-NOVUS, we have helped definitively reclassify 21 variants in the Qatari population.

## Discussion

We have attempted to help tackle India’s heavy cancer burden by establishing the variant pathogenicity landscape of all *BRCA1* and *BRCA2* gene variants in the Indian population reported in the IndiGen data through gold-standard ACMG/AMP guidelines. Using the pathogenic variants thus obtained, we have also established the genetic epidemiology of BRCA-linked HBOC in India. While the prevalence seems higher than established prevalence figures in other populations, it is likely because of several factors; a lack of uniform workflows in determining the pathogenicity of variants, even after the introduction of ACMG guidelines is still largely lacking, leading to potentially inaccurate classification of some variants. Further, with Next-generation sequencing becoming increasingly accessible even in clinical settings, more variants than ever before are being reported, and must be included in prevalence estimates. Another major problem is that a vast number of BRCA variants largely remain VUS. For example, of the 30,022 BRCA variants we queried from ClinVar, 49.2% were VUS. If we add all variants obtained from across the four databases, the number of ClinVar VUS variants increases to 96%. Since the BRCA genes are important to the study of breast cancer, it becomes essential that even the rare variants be classified for and an accurate prevalence be established. Another reason could be the fact that most prevalence estimates have been established before the advent of the ACMG guidelines. All these factors point towards the need for concerted efforts towards studying and establishing variant pathogenicity in greater detail.

The large number of VUS variants also makes it practically impossible to functionally classify all variants, or apply ACMG guidelines to them in a realistic timeline. We have therefore generated predictions for all non-benign variants using the brca-NOVUS tool. These classifications can act as a ready reference for clinicians and researchers studying genetic variants in the BRCA genes. They can further be utilized for estimating the genetic prevalence across different populations.

## Data Availability

All data produced in the present work are contained in the manuscript.
brca-NOVUS can be accessed at: https://github.com/aastha-v/brca-NOVUS

## Acknowledgements

Authors acknowledge funding from the Council of Scientific and Industrial Research (CSIR) through CNP-0007 Grant. The funders had no role in the preparation of the manuscript or decision to publish.

## Author Contributions

VS conceptualized, designed and supervised the study.

AV performed the analysis and complied the manuscript.

RCB, MI, VS, MKD, AM and BJ were involved in the generation of the study data.

VS and SSB conceived and designed the project. All authors have read and approved the final manuscript.

## Data Sharing And Code Availability

All data produced in the present work are contained in the manuscript.

brca-NOVUS can be accessed at: https://github.com/aastha-v/brca-NOVUS

