## Supplementary Data 1 for "Understanding the genetic epidemiology of hereditary breast cancer in India using whole genome data from 1029 healthy individuals"

**Supplementary Data 1. A detailed description of the methodology adopted for the annotation of ACMG-AMP guidelines.**

**Determination of PVS1**

PVS1 was marked for frameshift, stopgain or splice site variants using the AutoPVS1 tool. The tool is based on guidelines developed as part of the ClinGen Sequence Variant Interpretation (SVI) Workgroup's refined criteria for assigning the PVS1 attribute.
Only variants with "Strong" or "Very Strong" classification by the tool were assigned PVS1, and "Moderate" and "Supporting" were not considered.

**Determination of PS1 and PM5**

For determining PS1 and PM5 we matched the amino acid change of nonsynonymous variants with the pathogenic and likely pathogenic variants of ClinVar. If another ClinVar pathogenic variant had the same amino acid change as well as position at the protein level as our variant, but differed at the genomic level, our variant was marked PS1. In case of novel nonsynonymous variants , if the amino acid positions were the same at the protein level but the amino acid changes were different, PM5 was marked.

**Determination of PS2 and PM6**

Through extensive literature mining, we analyzed pedigrees: if a new mutation in the offspring was absent in the parents (de-novo mode of inheritance) then it was marked PS2. In case, the pedigree or the information did not sufficiently confirm the genotype of the parents, but the variant was assumed to be de-novo, then it was regarded as PM6.

**Determination of PS3 and BS3**

These attributes were annotated through literature screening to evaluate in vivo and in vitro functional studies that offered compelling evidence to establish the pathogenic or benign nature of a variant. If the variant was reported to be causative, it was marked PS3, while if it produced a benign effect, it was marked BS3.

**Determination of PS4**

This attribute was determined by the literature screening of case-control studies to determine the Odds Ratio (OR). If the OR was found to be greater than 5 and the Confidence interval (CI) was more than 1, then the variant was reported as PS4.

**Determination of PP1, PP1-M, PP1-S, and BS4**

We performed literature screening to analyze the co-segregation of the variant with the disease within a family. If the variant segregated with the phenotype in at least 3 individuals across at least 2 generations, PP1 was marked. The strength of the PP1 attribute depended on the number of diseased individuals the variant had segregated with, i.e. strong (PP1-S), moderate PP1-M), and supporting (PP1). If the variant was absent in any of the affected family members, or present in an unaffected family member, then due to lack of segregation, BS4 was assigned to the variant.

### Determination of PP2 and BP1

We calculated the total number of pathogenic missense and stop gain variants for each gene using Clinvar: if the percentage of missense variants in the gene was >80% and stop-gain variants was <20% then for nonsynonymous variants PP2 was marked. If stop-gain variants in the gene were more than 80% and nonsynonymous variant were < 20%, then the nonsynonymous variant was marked BP1.

### Determination of PP3 and BP4

The variants were annotated using the ljb26_all database of the ANNOVAR tool to determine the PP3 and BP4 parameters. The annotations included pathogenicity predictions from the benchmarked tools SIFT, PolyPhen2, and CADD. While SIFT cut-offs classify the variants as Deleterious (D) or Tolerated (T), PolyPhen2 predictions classify them into 3 categories as Probably Damaging (D), Possibly Damaging (P), or Benign (B). The CADD scores consist of PHRED-like scaled C-scores. We considered CADD scores above 15 to be interpreted as deleterious in our analysis. The variants were marked PP3 (pathogenic) if they were predicted to be deleterious by at least two of the three tools, or BP4 (benign) if a majority of the tools predicted them to be benign/tolerated.

### Determination of PP4

As the clinical sensitivity of testing could not be established as high (most patients testing positive for a pathogenic variant in that gene), and given that Hereditary Breast and Ovarian Cancer Syndrome (HBOC) often overlaps with other breast cancer syndromes, PP4 attribute was not awarded.

### Determination of PP5 and BP6

Variants were marked PP5 and BP6 using ClinVar (version 2020-01-13). Variants which had non-conflicting pathogenic/likely pathogenic calls were marked PP5, while those with non-conflicting benign/likely benign calls were marked BP6.

### Determination of PM1

We obtained the coordinates of protein domains in each gene as reported by Pfam using the Table Browser utility of UCSC Genome Browser. We intersected these with the coordinates of each of the variants: if the mutation laid within these domains, it was assigned the PM1 attribute.

**Determination of PM2, BA1, BS1, and BS2**

Allele frequencies from the control population datasets 1000 Genome Project (ALL.sites.2015_08), Exome Sequencing Project (esp6500siv2_all), and gnomADv3 were obtained from ANNOVAR to annotate the variants as BA1, BS1, and PM2. All variants with MAF> 5% in any of the three population datasets were marked BA1 while variants with MAF between 1 and 5% were considered as strong evidence to be benign for Mendelian disorders and marked BS1. In case the variant was present at an extremely low frequency (< 0.05%), it was classified as moderate evidence to be pathogenic (PM2). All variants that occurred in genes at greater than 1% frequency in the IndiGen dataset were marked BS2.

**Determination of PM3**

Since HBOC is inherited in an autosomal dominant manner with incomplete penetrance, we performed literature screening to check whether a second mutation had been detected along with our variant of interest. The presence of our variant in *cis* with a pathogenic variant was considered supporting evidence for a benign impact (BP2).

**Determination of PM4 and BP3**

For annotating in-frame insertions or deletions by overlapping them with the repeated region in the human genome using the repeat masker utility of the UCSC Genome Browser. If the in-frame insertions or deletions fell within the repeated region they were marked BP3, otherwise, they were marked PM4.

**Determination of BP7**

Synonymous variants not present within the splice site were marked BP7.

The attributes PS1, PS2, BP1, BP3, BP5, PM3, PM4, PM6, PP2, and PP4 were not assigned to our variants due to lack of sufficient evidence.
